# Neural correlates of Obsessive – Compulsive Personality Traits in Juvenile Myoclonic Epilepsy

**DOI:** 10.64898/2026.02.08.26345881

**Authors:** Lucas J Rainer, Bernardo Crespo Pimentel, Eugen Trinka, Giorgi Kuchukhidze, Mario Braun, Martin Kronbichler, Patrick Langthaler, Kornelius Winds, Georg Zimmermann, Lisa Kronbichler, Andreas Kaiser, Elisabeth Schmid, Elias Legat, Sarah Said-Yürekli, Aljoscha Thomschewski, Julia Höfler

## Abstract

**Objective:** To delineate the phenotype of juvenile myoclonic epilepsy (JME) with a focus on obsessive–compulsive personality disorder (OCPD) using multimodal psychiatric, neuropsychological, quantitative EEG (qEEG), and structural MRI markers within a predictive-processing/free-energy framework.

**Methods:** We prospectively studied 65 patients with JME and 68 matched healthy controls (HC). Participants completed DSM-IV SCID I/II interviews and a neuropsychological battery assessing working memory, psychomotor speed, mental flexibility, divided attention, inhibition, and phasic/tonic alertness; standard EEG and high-resolution structural MRI were acquired. Groups comprised HC and JME subgroups without psychiatric comorbidity, with non-OCPD Axis I/II diagnoses, and with OCPD. Welch’s t-tests (FDR-corrected) and Hedges’ g quantified neuropsychological and alpha-band coherence differences. Surface-based analyses assessed cortical thickness/surface area. Exploratory regressions tested associations of OCPD, seizure freedom, and antiseizure medication (ASM) load with cognition; Kendall’s tau tested coherence–cognition associations.

**Results:** Compared with HC, JME showed broad executive–attentional impairment, most pronounced in patients with psychiatric comorbidity. The OCPD subgroup exhibited particularly large slowing in psychomotor speed, inhibition (reaction time), and tonic alertness versus HC, while OCPD versus non-OCPD JME differences did not survive multiple-comparison correction. qEEG showed increased interhemispheric frontal and decreased temporal alpha coherence in JME, with temporal hypo-coherence strongest in those with psychiatric comorbidity; within JME, OCPD was linked to increased left fronto-temporal alpha coherence. In the MRI subsample, JME-OCPD demonstrated increased cortical thickness in left medial orbitofrontal and anterior cingulate regions (vs HC and vs JME without OCPD) and additional posterior occipito-temporal clusters versus HC. Regression and coherence–cognition associations were weak and non-significant after FDR correction.

**Significance:** JME features syndrome-level executive–attentional dysfunction and altered fronto–temporal network organization. Comorbid OCPD marks a subgroup with accentuated cognitive slowing and distinct medial prefrontal/cingulate structural and left fronto-temporal connectivity signatures, aligning with predictive-processing accounts of rigid, over-precise high-level priors.

**Key points:** JME is linked to broad executive–attentional impairment versus healthy controls. Psychiatric comorbidity amplifies cognitive deficits in JME.

JME with OCPD shows particularly large slowing/inhibitory-control deficits versus controls, while OCPD vs non-OCPD differences within JME are modest.

Alpha-band EEG coherence indicates altered network organization in JME and an OCPD-related increase in left fronto–temporal coherence within JME

Surface-based MRI suggests an OCPD-related structural phenotype in JME, involving medial orbitofrontal/anterior cingulate cortical thickening

## 1. Introduction

Juvenile myoclonic epilepsy (JME) is an idiopathic generalized epilepsy (IGE) syndrome, characterized by myoclonic jerks, generalized tonic-clonic seizures, and, in some individuals, absence seizures typically beginning in adolescence. ^1^ Although traditionally viewed as an epilepsy with preserved global cognition, growing evidence suggests that JME is associated with a more complex phenotype, including increased rates of psychiatric comorbidities and subtle but consistent impairments in executive functioning, decision-making, and social cognition.^2–4^ Some of these alterations are also present in unaffected relatives, supporting the view of JME as a disorder with inheritable neurobehavioral traits.^5,6^

Historical descriptions by Dieter Janz already highlighted a characteristic behavioral style in JME—combining reliability and diligence with impulsivity, affective lability, and difficulties with self-control, along with meticulousness, rule adherence, overcontrolled behavior, and reduced flexibility.^7–9^ This constellation, later summarized as the “Janz personality,” bears a striking resemblance to obsessive–compulsive personality disorder (OCPD), which is defined by perfectionism, cognitive rigidity, and excessive behavioral control.^10^ Nonetheless, the relationship between JME and obsessive–compulsive spectrum personality traits—as well as the associated neurobiological substrates—remains insufficiently understood.

Imaging studies in JME have reported alterations in large-scale networks, including abnormal frontal connectivity on EEG and structural and functional abnormalities in frontal and temporal cortices.^11–14^ Most prior work, however, has focused on single modalities and has rarely integrated psychiatric, neuropsychological, EEG, and structural MRI data within the same cohort. As a result, it is unclear whether the so-called “Janz personality” reflects a specific personality phenotype, a consequence of seizures or a manifestation of the underlying neurodevelopmental pathophysiology.

The Free Energy Principle conceptualizes neurological and psychiatric disorders within predictive-processing and free-energy frameworks, in which the brain is seen as a hierarchical generative model that minimizes prediction error by updating internal beliefs and sampling the environment.^15,16^ Within this view, executive dysfunction, altered fronto–temporal connectivity, and rigid personality traits may reflect aberrant precision-weighting of prediction errors and over-stabilization of high-level priors about control and order, leading to psychiatric, as well as neurologic disorders.

In this multimodal study, we combine structured psychiatric assessment, neuropsychological testing, quantitative EEG (qEEG), and surface-based MRI (sMRI) in patients with JME and matched healthy controls. Specifically, we hypothesize that comorbid OCPD marks a distinct endophenotype of JME and interpret our findings in the light of predictive-processing and free-energy accounts.

## 2. Materials and Methods

### 2.1.1 Participants

We prospectively recruited 65 patients with JME between February 2017 and December 2020 at the Department of Neurology, Christian Doppler University Hospital, Paracelsus Medical University, Salzburg, Austria, and 68 healthy controls matched for age, sex, and education. JME was diagnosed according to International League Against Epilepsy criteria.^1^ Exclusion criteria for patients were neurological illnesses other than JME, structural lesions on MRI, seizures <72 hours before MRI, pregnancy, and acute benzodiazepine use. Controls were excluded if they had a history of neurological or psychiatric disorders, known structural brain lesions, MRI incompatibility, or pregnancy.

### 2.1.2 Standard Protocol Approvals, Registrations and Patient Consents

All participants gave written informed consent; the study was approved by the local ethics committee (No. E1638) and conducted in accordance with the Declaration of Helsinki. All participants provided written informed consent.

### 2.2 Procedure

All participants underwent structured psychiatric and neuropsychological assessment, standard EEG, and structural MRI.

#### 2.2.1 Neuropsychological & psychiatric Assessment

Neuropsychological testing focused on attention and executive functions. We used three subtests of the Test of Attentional Performance (TAP; Alertness, Divided Attention, Go/NoGo)^17^ to assess intrinsic and phasic alertness, shared auditory–visual attention, and inhibitory control; the Trail Making Test (TMT-A/B)^18^ to index psychomotor speed and mental flexibility; and the Digit Span subtests (forward and backward) of the Wechsler Adult Intelligence Scale IV^19^ to assess verbal attention span and working memory.

Psychiatric diagnoses were obtained using the German versions of the Structured Clinical Interview for DSM-IV Axis I (SCID I) and Axis II (SCID II)^20,21^, which are considered gold-standard semi-structured interviews for clinical and personality disorders. OCPD status and other Axis II diagnoses were derived from SCID II. Detailed test procedures are provided in the Supplementary Methods S1.

#### 2.2.2 EEG acquisition and preprocessing

EEG was recorded for approximately 25 minutes using 21 scalp electrodes placed according to the international 10–20 system, referenced to Cz, with impedances <5 kΩ and a sampling rate of 256 Hz. Standard procedures included resting with eyes closed, occipital alpha suppression testing, photic stimulation and a hyperventilation period (if not contraindicated). For the present analyses, we focused on artifact-free resting segments during hyperventilation, as these epochs were least contaminated by movement and were deemed most sensitive for capturing epilepsy-related network disturbances. Six patients were excluded due to poor signal quality or absence of a hyperventilation period. Signals were re-referenced to the common average (excluding Cz, A1, A2), and analyses were restricted to frontal and temporal channels (Fp1, F3, F7, Fp2, F4, F8, T3, T5, T4, T6). EEGs were segmented into a 3-minute window starting at the onset of hyperventilation.

Multivariate autoregressive models were estimated for each participant, and alpha-band (8–13 Hz) connectivity was derived using real-valued coherence^22^ as an undirected measure. To limit the number of tests, only alpha-band coherence values from six a priori defined fronto–temporal region-of-interest (ROI) connections were entered into group analyses (frontal left to frontal right; frontal left to temporal left; frontal right to temporal right; temporal right to temporal left). Full modelling details are provided in the Supplementary Methods S2.

#### 2.2.3 MRI acquisition and surface-based metrics

Structural MRI was acquired on a 3T Magnetom Prisma-Fit scanner using a 3D multiecho MPRAGE sequence (voxel size 0.8 × 0.8 × 0.8 mm³). Standard FreeSurfer pipelines were used for cortical reconstruction, including skull stripping, intensity normalization, segmentation, and surface extraction (∼16,000 vertices per hemisphere). Surfaces were visually inspected and manually corrected, when necessary, then registered to a common template.

We focused on cortical surface markers—cortical thickness, white matter surface area, and local gyrification index (LGI)—to capture three-dimensional properties of the cortex. Cortical volume was not analyzed separately because it is a derived measure of thickness and surface area. Surface area was computed as the average area of the six triangles surrounding each vertex on the white matter surface; LGI was computed using the standard FreeSurfer implementation. Cortical thickness and surface area maps were smoothed with a 20-mm kernel, LGI with 5 mm due to inherent smoothing. Detailed parameters are provided in the Supplement S3.

### 2.3. Statistical analysis

#### 2.3.1 Clinical and neuropsychological data

All statistical analyses were performed in IBM SPSS Statistics (IBM Corp.), except for the vertexwise surface-based analyses, which were conducted using BrainStat for MATLAB^23^. Demographic and clinical variables were compared between JME and controls using *χ²* tests or non-parametric tests, as appropriate. For neuropsychological outcomes, we computed descriptive statistics and compared groups using Welch’s *t*-tests (two-tailed, unequal variances) for a priori contrasts: (1) JME with OCPD vs healthy controls, (2) JME with OCPD vs JME without OCPD.

Additional exploratory contrasts (all JME vs controls; JME with any psychiatric disorder vs controls) are reported in the Supplement Table S1. Given multiple neuropsychological measures, *p*-values were corrected within each contrast using the Benjamini–Hochberg false discovery rate (FDR). We report FDR-corrected *p*-values (*p_FDR_*) and Hedges’ *g* as effect size.

#### 2.3.2 EEG alpha-band coherence

For each of the six ROI connections, alpha-band coherence was compared between groups defined by psychiatric status (JME without psychiatric comorbidity, JME with non-OCPD psychiatric diagnoses, JME with OCPD, healthy controls) using Welch’s t-tests for four contrasts: (1) all JME vs controls, (2) JME with any psychiatric comorbidity vs controls, (3) JME with OCPD vs controls, and (4) JME with OCPD vs JME without psychiatric comorbidity. FDR correction across the six connections was applied per contrast.

#### 2.3.3 Surface-Based Exploratory Analysis

Vertexwise analyses were performed using BrainStat for MATLAB. Age effects were first modeled in controls and then incorporated into group comparisons as covariates, together with sex and, for surface area, total white matter volume. Cluster-wise family-wise error (FWE) correction was applied with a cluster-defining threshold of *p* < 0.025.

#### 2.3.4 Exploratory models and correlations

To explore clinical determinants of cognition, we computed separate linear regression models, where each neuropsychological outcome was fed as dependent variable, and OCPD status, seizure freedom, and total antiseizure medication (ASM) load as predictors. FDR correction across outcomes was applied per predictor.

To examine associations between alpha coherence and cognition across all participants, Kendall’s tau correlations were computed between alpha coherence in each connection and six cognitive measures (Go/NoGo reaction time, TMT-B, digit span forward/backward, verbal learning, divided-attention errors), with FDR correction across all correlations. Full results are reported in the Supplement Tables S2 and S3.

## 3. Results

### 3.1. Clinical and demographic data

Sixty-five patients with JME and 68 healthy controls were included. Groups did not differ significantly in median age (controls: 24.0 years, IQR 21.0–31.5; JME: 27.0 years, IQR 23.0–34.0; p = .171) or sex distribution (women: 54% vs 58%; p = .727). Years of education were comparable, with a trend toward slightly fewer years in JME (controls: median 12.0, IQR 12.0–15.0; JME: median 12.0, IQR 11.0–14.0; p = .052). In JME, median age at onset was 14.0 years (IQR 12.0–16.0), and median epilepsy duration 14.0 years (IQR 8.0–19.0). At study time, 30.8% had generalized tonic–clonic seizures, 25% absences, 57% myoclonic seizures, and 38% reported no seizures. Five patients (8%) were off medication, 58% on monotherapy, and 34% on polytherapy; levetiracetam (61%), valproic acid (29%), and lamotrigine (17%) were the most common ASM, and 17% received antidepressants (Table 1).

**Table 1.**
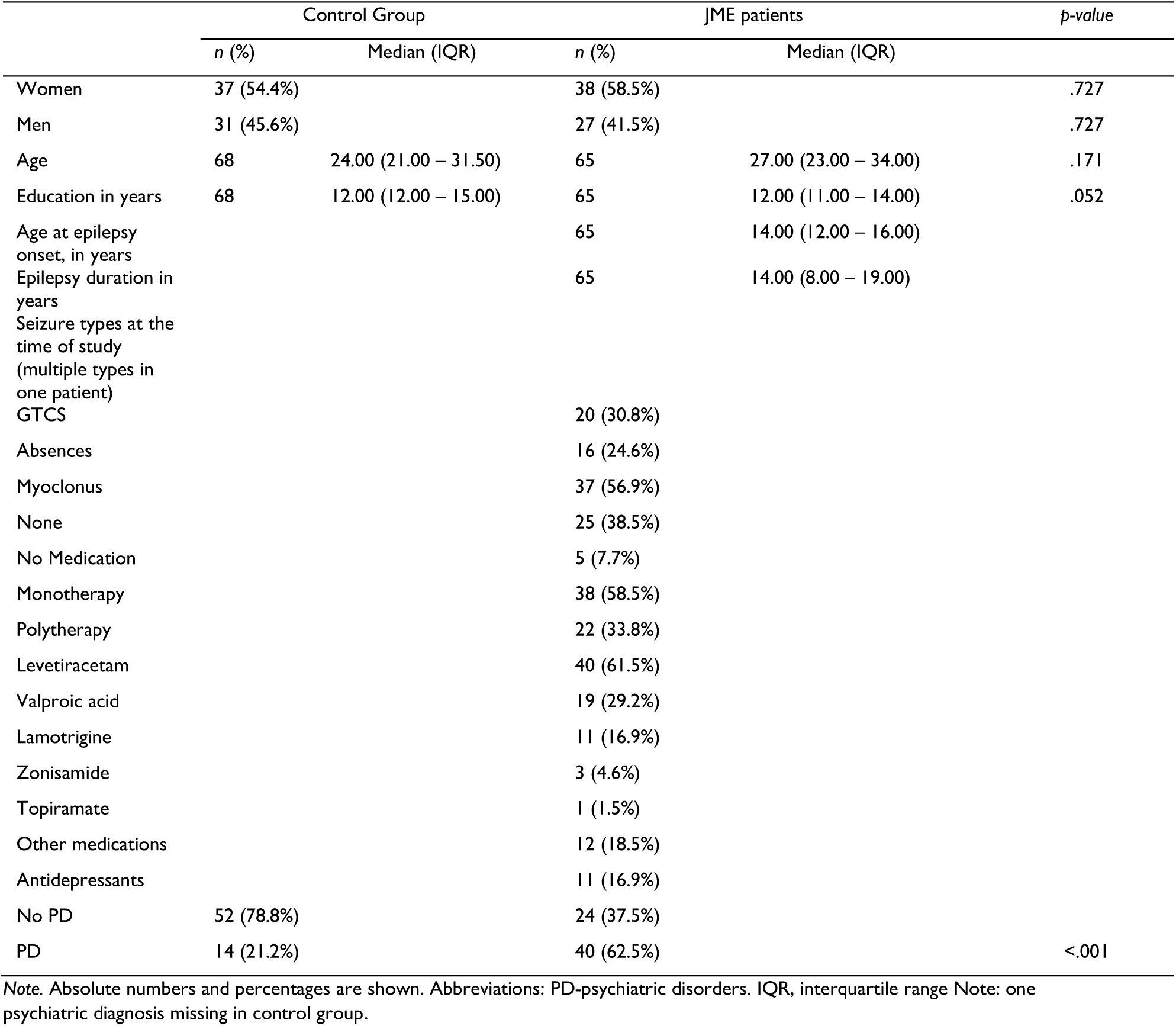
Demographic and clinical features.

|  | Control Group |  | JME patients |  | p-value |
| --- | --- | --- | --- | --- | --- |
|  | n (%) | Median (IQR) | n (%) | Median (IQR) |  |
| Women | 37 (54.4%) |  | 38 (58.5%) |  | .727 |
| Men | 31 (45.6%) |  | 27 (41.5%) |  | .727 |
| Age | 68 | 24.00 (21.00 – 31.50) | 65 | 27.00 (23.00 – 34.00) | .171 |
| Education in years | 68 | 12.00 (12.00 – 15.00) | 65 | 12.00 (11.00 – 14.00) | .052 |
| Age at epilepsy onset, in years |  |  | 65 | 14.00 (12.00 – 16.00) |  |
| Epilepsy duration in years |  |  | 65 | 14.00 (8.00 – 19.00) |  |
| Seizure types at the time of study (multiple types in one patient) |  |  |  |  |  |
| GTCS |  |  | 20 (30.8%) |  |  |
| Absences |  |  | 16 (24.6%) |  |  |
| Myoclonus |  |  | 37 (56.9%) |  |  |
| None |  |  | 25 (38.5%) |  |  |
| No Medication |  |  | 5 (7.7%) |  |  |
| Monotherapy |  |  | 38 (58.5%) |  |  |
| Polytherapy |  |  | 22 (33.8%) |  |  |
| Levetiracetam |  |  | 40 (61.5%) |  |  |
| Valproic acid |  |  | 19 (29.2%) |  |  |
| Lamotrigine |  |  | 11 (16.9%) |  |  |
| Zonisamide |  |  | 3 (4.6%) |  |  |
| Topiramate |  |  | 1 (1.5%) |  |  |
| Other medications |  |  | 12 (18.5%) |  |  |
| Antidepressants |  |  | 11 (16.9%) |  |  |
| No PD | 52 (78.8%) |  | 24 (37.5%) |  |  |
| PD | 14 (21.2%) |  | 40 (62.5%) |  | <.001 |

Note. Absolute numbers and percentages are shown. Abbreviations: PD-psychiatric disorders. IQR, interquartile range Note: one psychiatric diagnosis missing in control group.

### 3.2 Psychiatric comorbidity

Among participants with complete SCID data (66 controls, 64 JME), any Axis I and/or II disorder was diagnosed in 62% (40/64) of JME patients versus 21% (14/66) of controls (*p* < .001). Axis I disorders alone did not differ significantly between groups (17% vs 12%; *p* = .464), whereas Axis II disorders were markedly more frequent in JME—both when present without Axis I pathology (27% vs 6%; *p* = .002) and when combined with Axis I disorders (19% vs 3%; *p* = .004). Histrionic personality disorder occurred exclusively in JME (8% vs 0%; *p* = .027). OCPD was present in 22% (14/64) of JME patients and 3% (2/66) of controls (*p* = .001; Table 2).

**Table 2.** Descriptive statistics of psychiatric comorbidities.

| Axes | Control Group<br>(n=66) | JME<br>patients<br>(n=64) | p-value |
| --- | --- | --- | --- |
|  | n (%) |  |  |
| With psychiatric disorder | 14 (21.2%) | 40 (62.5%) | <.001 |
| Axis I disorder only | 8 (12.1%) | 11 (17.2%) | .464 |
| Axis II disorder only | 4 (6.1%) | 17 (26.6%) | .002 |
| Axis I & II disorder | 2 (3.0%) | 12 (18.8%) | .004 |
| *Current DSM-IV diagnoses, multiple diagnoses in one patient* |  |  |  |
| *Axis I disorders* |  |  |  |
| Affective disorders (296.2x, 296.3x, 293.83) | 5 (7.6%) | 10 (15.6%) | .178 |
| Schizophrenia and other psychotic disorders (298.8, 295.30, 293.81) | 1 (1.5%) | 2 (3.1%) | .616 |
| Substance-related disorders (303.90, 304.30, 304.80) | 5 (7.6%) | 7 (10.9%) | .558 |
| Anxiety disorders (300.02, 300.21, 300.01, 300.22, 300.29, 300.23, 300.3, 309.81, 308.3) | 2 (3%) | 8 (12.5%) | .053 |
| Eating disorders (307.1, 307.51, 307.50) | 0 (0%) | 3 (4.7%) | .117 |
| Adjustment disorders (309.9, 309.24, 309.0) | 4 (6.1%) | 5 (7.8%) | .742 |
| *Axis II disorders* |  |  |  |
| *Cluster A* |  |  |  |
| Paranoid personality disorder (301.00) | 1 (1.5%) | 4 (6.2%) | .204 |
| Schizotypal personality disorder (301.22) | 1 (1.5%) | 0 (0%) | 1 |
| *Cluster B* |  |  |  |
| Antisocial personality disorder (301.7) | 2 (3%) | 5 (7.8%) | .270 |
| Borderline personality disorder (301.83) | 3 (4.5%) | 5 (7.8%) | .489 |
| Histrionic personality disorder (301.50) | 0 (0%) | 5 (7.8%) | .027 |
| Narcissistic personality disorder (301.81) | 1 (1.5%) | 4 (6.2%) | .204 |
| *Cluster C* |  |  |  |
| Avoidant personality disorder (301.82) | 1 (1.5%) | 5 (7.8%) | .112 |
| Dependent personality disorder (301.83) | 1 (1.5%) | 3 (4.7%) | .362 |
| Obsessive-compulsive personality disorder (301.4) | 2 (3%) | 14 (21.9%) | .001 |
| *Personality disorder not otherwise specified* |  |  |  |
| with depressive personality traits (301.9) | 1 (1.5%) | 4 (6.2%) | .204 |
| with passive-aggressive personality traits (301.9) | 1 (1.5%) | 4 (6.2%) | .204 |

Note. Absolute numbers and percentages are shown. Abbreviations: JME, juvenile myoclonic epilepsy; DSM-IV, Diagnostic and Statistical Manual of Mental Disorders-IV

### 3.3 Neuropsychological performance

At the group level, JME patients showed broad impairments relative to controls (full results in Supplement Table S4). In the total JME group, digit span forward and backward, inhibition (reaction time), divided attention, mental flexibility, psychomotor speed, and phasic and tonic alertness were all significantly impaired after FDR correction, with effect sizes ranging from small to large.

In line with our primary focus, we examined subgroups defined by OCPD status. Descriptive data for healthy controls, JME without OCPD, and JME with OCPD are shown in Table 3.

**Table 3.** Neuropsychological performance in healthy controls, JME without OCPD, and JME with OCPD.

| Measure | HC (n) | HC M (SD) | JME non-OCPD<br>(n) | JME non-OCPD<br>M (SD) | JME OCPD (n) | JME OCPD M<br>(SD) |
| --- | --- | --- | --- | --- | --- | --- |
| Digit span (back) | 67 | 7.22 (1.91) | 48 | 6.29 (1.80) | 12 | 6.42 (2.27) |
| Digit span (fwd) | 67 | 10.31 (2.17) | 48 | 9.23 (2.36) | 12 | 9.50 (1.83) |
| Inhibition (err) | 65 | 1.52 (1.48) | 47 | 1.87 (1.84) | 13 | 2.00 (1.58) |
| Inhibition (rt) | 65 | 380.02 (48.18) | 47 | 426.04 (75.60) | 13 | 436.15 (45.51) |
| Divided attention (vis) | 66 | 788.24 (94.58) | 47 | 817.30 (127.17) | 13 | 893.54 (216.77) |
| Divided attention (aud) | 66 | 613.91 (79.05) | 47 | 637.17 (155.60) | 13 | 656.00 (86.61) |
| Mental flexibility | 67 | 51.82 (16.74) | 48 | 68.35 (40.00) | 12 | 77.42 (41.78) |
| Psychomotor speed | 67 | 22.19 (5.60) | 48 | 28.31 (9.29) | 12 | 31.58 (10.10) |
| Tonic alertness (rt) | 66 | 231.11 (24.15) | 47 | 259.21 (71.37) | 13 | 280.46 (50.72) |
| Phasic alertness (rt) | 66 | 230.42 (27.42) | 47 | 252.30 (64.15) | 13 | 265.69 (55.10) |

Note. Values are mean (M) and standard deviations (SD). Group sizes (n) vary across measures due to missing data. Higher values in reaction-time measures (inhibition RT, divided attention, psychomotor speed, tonic and phasic alertness) indicate slower performance. JME = juvenile myoclonic epilepsy; OCPD = obsessive–compulsive personality disorder; HC = healthy controls; err = errors; rt = reaction time; vis = visual modality; aud = auditory modality.

Patients with JME and OCPD (JME-OCPD) performed consistently worse than controls. After FDR correction, large and robust effects were observed for psychomotor speed, inhibition reaction time, and tonic alertness (Hedges’ *g* ≈ 1.16–1.64; all *p_FDR_* ≤ .011), indicating marked slowing and reduced alertness. For working memory, divided attention, mental flexibility, and phasic alertness, the OCPD group also tended to perform worse than controls, but these differences did not survive FDR correction and should be interpreted cautiously.

Direct comparisons between JME-OCPD and JME without OCPD did not yield statistically significant differences after FDR correction on any neuropsychological measure (all *p_FDR_* > .20), and effect sizes were generally small, with small-to-moderate effects for visual divided attention and psychomotor speed at most (Table S4). Overall, cognitive deficits in working memory, attention, and executive functioning that appear to characterize JME as a syndrome, are amplified in those with psychiatric comorbidity (including OCPD), but do not sharply separate JME patients with OCPD from those without. Figure 1 summarizes significant neuropsychological group differences in terms of Hedges’ *g*.

**Figure 1.**
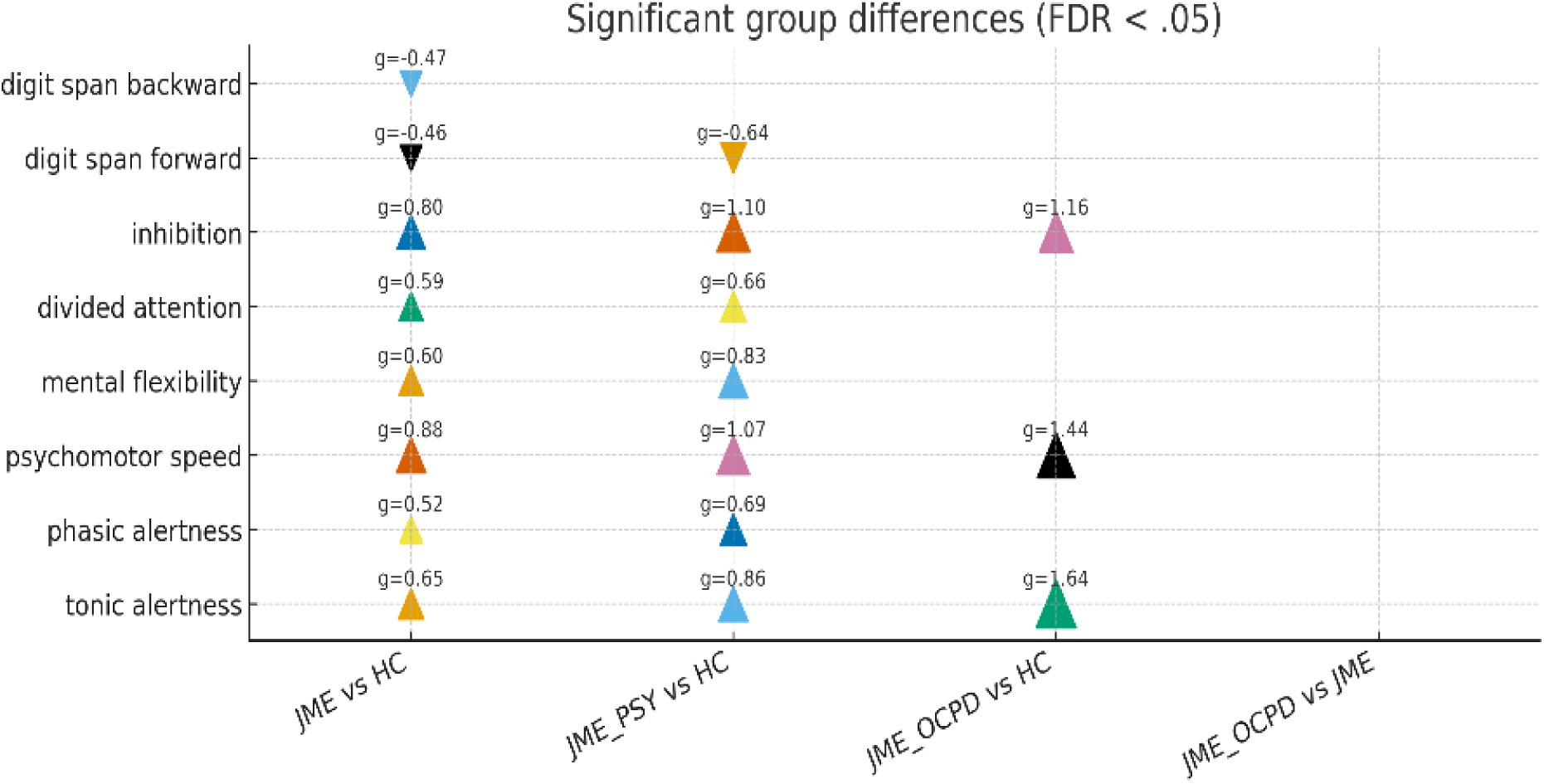
Significant neuropsychological group differences (FDR-corrected, *p* < .05) *Note.* Triangles depict effect sizes (Hedges’ *g*) for neuropsychological measures showing significant group differences after false discovery rate (FDR) correction (*p* < .05). Positive values indicate higher scores in the JME group relative to the comparison group, negative values indicate lower scores. JME = juvenile myoclonic epilepsy; HC = healthy controls; JME_PSY = JME with any Axis I and/or Axis II psychiatric disorder; JME_OCPD = JME with obsessive–compulsive personality disorder. Digit span backward/forward = WAIS-IV Digit Span subtests; inhibition = Go/NoGo reaction time; divided attention = TAP visual–auditory divided attention; psychomotor speed = TMT-A completion time; mental flexibility = TMT-B completion time; phasic/tonic alertness = TAP Alertness reaction times.

### 3.4 qEEG and Alpha-band coherence

Alpha-band coherence analyses focused on six a priori fronto–temporal connections in 52 JME patients (23 without psychiatric comorbidity, 15 with non-OCPD psychiatric diagnoses, 14 with OCPD) and 53 controls. Compared with controls, the JME group showed significantly increased interhemispheric frontal coherence in both directions (frontal left to frontal right and frontal right to frontal left; *t* ≈ 2.65, *p_FDR_* = .019, *g* ≈ 0.51) and significantly reduced coherence from right to left temporal regions (temporal right to temporal left; *t* = – 3.43, *p_FDR_* = .005, *g* = –0.66; Table 4). This pattern suggests frontal hyper-coherence and temporal hypo-coherence in the alpha band.

**Table 4.**
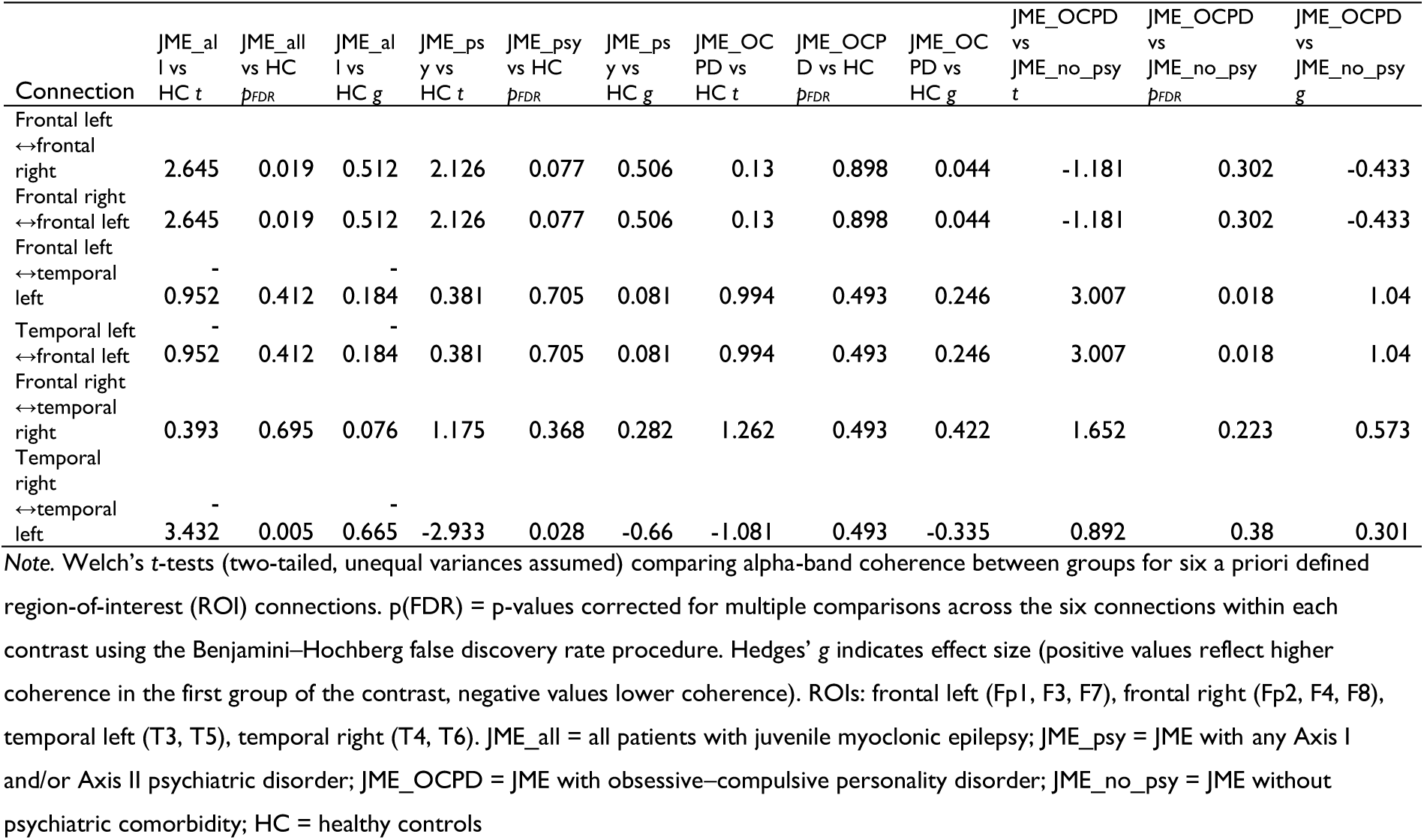
Group differences in alpha-band coherence between frontal and temporal regions.

In JME patients with any psychiatric comorbidity, temporal alpha hypo-coherence (temporal right to temporal left) remained significant versus controls (*t* ≈ –2.93, *p_FDR_* = .028, *g* ≈ –0.66), whereas frontal hyper-coherence reached only trend level (*p_FDR_* ≈ .08). JME-OCPD did not differ significantly from controls in any alpha connection after FDR correction (all *p_FDR_* ≥ .49).

When JME-OCPD participants were compared directly with JME patients without psychiatric comorbidity, a robust increase in left fronto–temporal coherence emerged: coherence between frontal left and temporal left regions was significantly higher in both directions (*t* ≈ 3.01, *p_FDR_* = .018, *g* ≈ 1.04). No other alpha-band connections differed significantly between these JME subgroups. Taken together, our JME cohort is characterized by increased frontal and reduced temporal interhemispheric alpha coherence, temporal hypo-coherence is most pronounced in individuals with psychiatric comorbidity, and comorbid OCPD is associated with abnormally elevated left fronto–temporal coherence within the JME group.

### 3.5 Structural MRI

In the structural MRI subsample, surface-based vertexwise analyses revealed focal increases in cortical thickness in patients with JME and OCPD. Compared with healthy controls, the JME-OCPD group showed significant clusters of increased cortical thickness in the left medial orbitofrontal cortex (*p_FWE_* < .01) and the left anterior cingulate cortex (*p_FWE_* < .05). In addition, significant thickness increases were observed in right occipital/ventral visual regions including the lingual cortex (*p_FWE_* < .05), fusiform cortex (*p_FWE_* < .05), and pericalcarine cortex (*p_FWE_* < .01). No clusters showed reduced cortical thickness in JME-OCPD compared with controls at the applied threshold.

No clusters survived FWE correction in the comparison of JME without OCPD versus healthy controls.

When directly comparing JME-OCPD with JME without OCPD, the JME-OCPD group again showed increased cortical thickness in the left medial orbitofrontal cortex (*p_FWE_* < .01) and the left anterior cingulate cortex (*p_FWE_* < .05). Please see figure 2, A and B, as well as table S5 in the Supplementary for detailed information.

**Figure 2.**
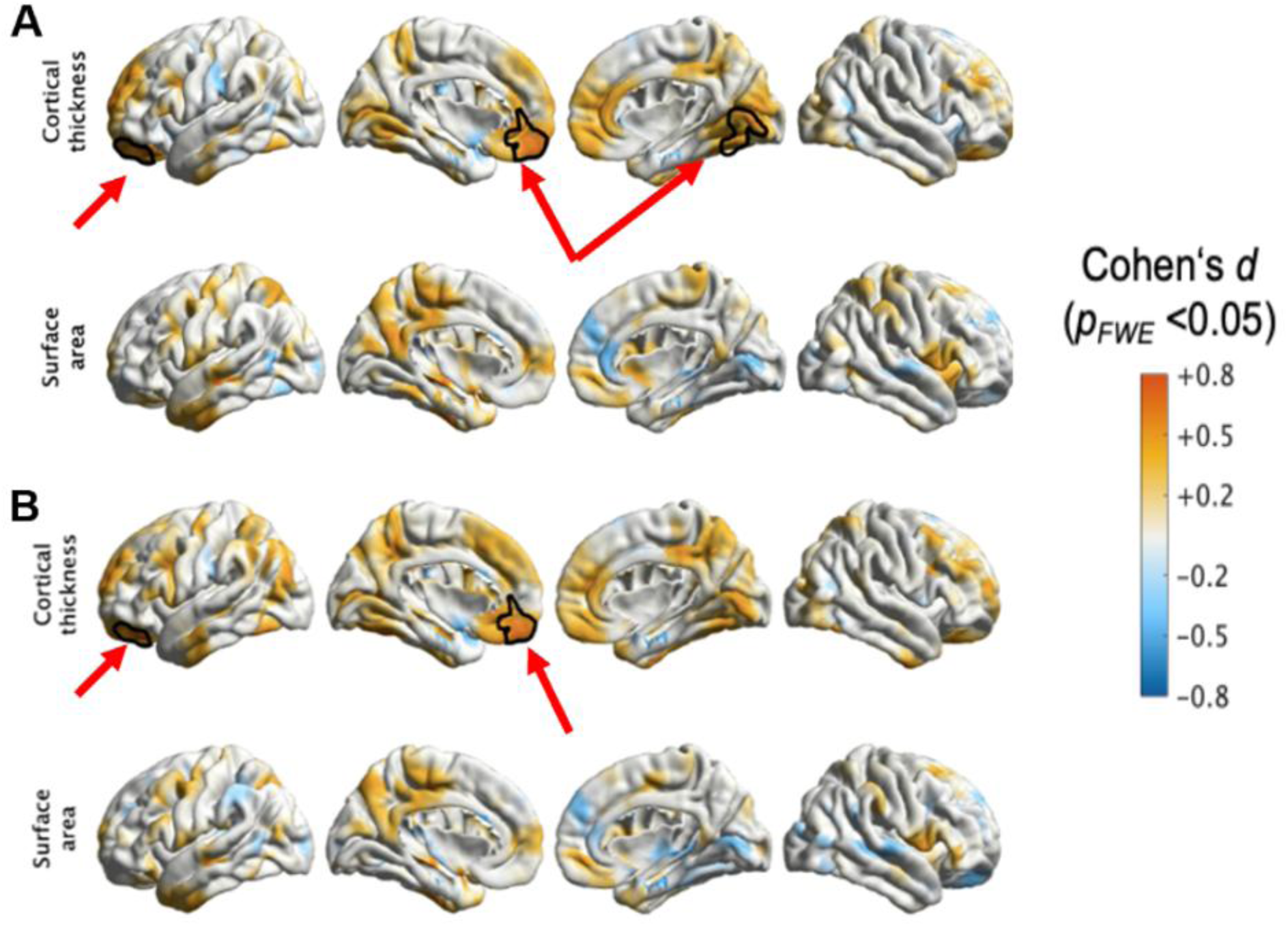
Increased cortical thickness in JME with OCPD compared with healthy controls (A) and all other JME patients (B). *Note.* Surface-based vertexwise comparisons of cortical thickness between JME patients with OCPD and healthy controls (A) and between JME patients with OCPD and JME patients without OCPD (B). **A:** Black outlines indicate clusters with significantly higher cortical thickness in JME-OCPD than in healthy controls in the left medial orbitofrontal cortex, left anterior cingulate cortex, and right occipital/ventral visual regions (lingual, fusiform, and pericalcarine cortex) (cluster-wise *pFWE* < .05; cluster-defining threshold *p* < .025). **B:** Black outlines indicate clusters with significantly higher cortical thickness in JME-OCPD than in JME without OCPD in the left medial orbitofrontal cortex and left anterior cingulate cortex (cluster-wise *pFWE* < .05; cluster-defining threshold *p* < .025). No clusters showed significantly reduced cortical thickness at this threshold. Maps depict effect sizes (Cohen’s d); warm colors indicate higher values in JME-OCPD relative to the comparison group, cool colors indicate lower values.

### 3.6 Exploratory regression and coherence–cognition associations

Exploratory linear regression models relating neuropsychological performance to OCPD status, seizure freedom, and total ASM load explained only a small proportion of variance across measures (*R²* ≤ .17; Table S2, Supplement). No predictor remained significant after FDR correction (all *p_FDR_* ≥ .33).

Across all participants, Kendall’s tau correlations between alpha-band coherence and cognitive performance were weak (|*τ*| ≤ .18), and none survived FDR correction (all *p_FDR_* = 1.00; Table S3, Supplement). Uncorrected trends suggested slightly slower Go/NoGo responses with higher alpha coherence in selected fronto–temporal connections, but no consistent pattern emerged. Overall, these exploratory analyses provide no robust evidence that resting-state alpha coherence during hyperventilation is strongly coupled to standard neuropsychological test performance.

## 4. Discussion

Across modalities, our findings converge on fronto-temporal networks that support executive control, attentional regulation, and personality-related functions. At the syndrome level, JME is associated with broad executive–attentional deficits and an imbalance between frontal and temporal resting-state connectivity. Within this framework, psychiatric comorbidity, and OCPD in particular, identifies a distinct endophenotype with more severe cognitive impairment and distinct neurobiological features, including left fronto-temporal alpha hyper-coherence and increased cortical thickness in left medial orbitofrontal, left anterior cingulate cortex, right lingual/fusiform/pericalcarine clusters compared to controls, as well as left medial orbitofrontal and left anterior cingulate cortex in JME with OCPD compared to JME without OCPD.

### Neuropsychological performance

Our findings support the view of juvenile myoclonic epilepsy (JME) as a broader neurodevelopmental condition with clinically meaningful cognitive sequelae. Across the JME cohort, performance was reduced relative to healthy controls in executive–attentional domains, including working memory, inhibition, processing speed, mental flexibility, divided attention, and tonic/phasic alertness. This pattern aligns with prior evidence that cognitive control and speeded processing are core vulnerabilities in JME rather than rare complications.^2,4^

Psychiatric comorbidity further shaped this cognitive profile. JME patients with any Axis I/II diagnosis showed the most consistent and pronounced impairments, suggesting that psychiatric burden amplifies an already vulnerable executive system. In the subgroup with obsessive–compulsive personality disorder (OCPD), deficits were particularly marked for psychomotor speed, inhibition reaction time, and tonic alertness when compared with healthy controls, whereas other measures showed directionally similar but less robust effects after correction. Importantly, however, direct comparisons between JME patients with and without OCPD did not yield FDR-significant differences, and effect sizes were small-to-moderate at most. Together with the exploratory regression models (low explained variance; no predictor surviving FDR correction), this argues against a simple one-to-one mapping between current clinical status (OCPD, seizure freedom, ASM load) and cognitive performance.^2^ A parsimonious interpretation is that executive–attentional dysfunction is a syndrome-level feature of JME that can be exacerbated by psychiatric comorbidity, including OCPD, but is not uniquely determined by it.

### EEG alpha-band coherence

The qEEG results add network-level evidence for altered large-scale organization in JME. When considering all JME patients, alpha-band coherence was increased between left and right frontal regions, whereas temporo-temporal coherence from right to left was reduced. Taken together, frontal hyper-coherence alongside reduced temporal interhemispheric coupling is compatible with an imbalance between control-related frontal systems and temporo-limbic integration during rest-like conditions.^24,25^; in predictive-processing accounts, alpha rhythms are thought to regulate the precision of prediction errors and gate information flow in hierarchical cortical networks.^26,27^

A key methodological feature is that alpha coherence was derived from resting epochs during the hyperventilation period. Clinically, hyperventilation is used to increase the yield of epileptiform activity, but it also constitutes a standardized physiological challenge that many participants experience as mildly stressful.^28^ Our coherence patterns therefore likely reflect not only baseline coupling but also network configuration under a controlled perturbation.

One speculative, but clinically guided, interpretation is that stress-related increases in frontal and fronto-temporal alpha synchrony may index a shift toward more rigid, overcontrolled network states.^29^ In everyday contexts, such a stress-linked “tightening” of control could contribute to inflexible, rule-bound behavior and, when control is exceeded, abrupt dysregulated responses—phenomena often noted clinically in JME.^30^

Within the broader JME pattern, OCPD was associated with a more circumscribed signature. JME-OCPD did not differ from healthy controls after FDR correction on the predefined alpha connections, suggesting that OCPD does not simply add a global increase or decrease in alpha coherence beyond the syndrome-level JME effects^13^

However, when contrasted with JME patients without any psychiatric comorbidity, JME-OCPD showed markedly increased left fronto-temporal coherence (bidirectionally) with a large effect size. This finding is consistent with the idea that OCPD traits in JME relate to a specific configuration of left-lateralized fronto-temporal coupling, potentially relevant to overcontrol and reduced flexibility described historically in the “JME personality” accounts.^7,9^

Finally, correlations between alpha coherence and cognition were small and did not survive multiple-comparison correction. This suggests that alpha coherence may index trait-like properties of network organization under mildly challenging conditions, that are only partially captured by performance on brief standardized tasks, especially in modest samples.

### Structural MRI findings

The structural MRI results converge with the EEG findings in implicating circuits supporting control and monitoring functions in JME patients with OCPD. Surface-based analyses showed increased cortical thickness in the medial orbitofrontal cortex and anterior cingulate cortex (ACC) in JME-OCPD compared with healthy controls, and the same medial prefrontal–cingulate pattern re-emerged when JME-OCPD was compared with JME without OCPD. Replication across contrasts strengthens the impression that medial orbitofrontal and ACC regions are specifically involved in the OCPD-related phenotype within JME rather than reflecting a general feature of JME.^31^

Functionally, medial orbitofrontal regions contribute to value-based evaluation, outcome appraisal, and regulation of behavior according to expected consequences,^32,33^ while the ACC supports performance monitoring, conflict detection, and the allocation of cognitive control—i.e., signaling when behavior should be tightened, corrected, or stabilized. ^34,35^ Orbitofrontal–cingulate thickening therefore fits a neurodevelopmental architecture biased toward over-monitoring and over-control, consistent with core OCPD traits (perfectionism, over-conscientiousness, intolerance of uncertainty).^36^

In addition, JME-OCPD differed from healthy controls by increased thickness in right lingual, fusiform, and pericalcarine cortex. These regions support early visual processing and ventral stream functions relevant to detailed visual analysis. While this posterior pattern should be interpreted cautiously, one possibility is that altered visual cortical architecture relates to enhanced weighting of perceptual detail under uncertainty, dovetailing with clinical tendencies toward meticulousness and checking-like control strategies.^37,38^ Notably, these posterior clusters did not re-appear in the within-JME contrast, suggesting they may reflect case–control differences that are most detectable when contrasting JME-OCPD with healthy controls.

### A free-energy perspective on JME and OCPD

The multimodal pattern, consisting of executive–attentional deficits at the syndrome level, alpha-band coherence abnormalities under a standardized perturbation, and medial prefrontal–cingulate thickening in JME-OCPD, can be integrated within predictive-processing and free-energy accounts.^39^ In these frameworks, cognition and control are framed as hierarchical inference: the system minimizes prediction error by updating beliefs and by selecting actions that sample evidence. A crucial mechanism is precision-weighting, i.e., how strongly prediction errors are trusted and propagated within cortical hierarchies.^15,40^

Alpha rhythms have been proposed as one physiological substrate contributing to precision control and long-range gating.^27^ From this perspective, frontal hyper-coherence together with reduced temporal interhemispheric coupling may reflect altered precision allocation across fronto-temporal systems: overly synchronized frontal networks may instantiate overly stable control priors, whereas reduced temporal coupling may limit contextual and affective integration. The fact that our alpha measures were derived during hyperventilation further suggests that these patterns capture how precision and coupling reconfigure under challenge rather than in a purely neutral resting state, in our perspective potentially relevant to real-life stress sensitivity.

Within this computational framing, OCPD can be conceptualized as excessive precision assigned to high-level beliefs about order, correctness, and threat avoidance, such that discrepant evidence is underweighted and behavior becomes rigid and overcontrolled. ^40^ Increased left fronto-temporal coherence, together with medial orbitofrontal–ACC thickening, may index a neurobiological architecture in which evaluative and monitoring systems exert strong top-down influence, biasing the organism toward control-tightening strategies to resolve uncertainty. This provides a plausible bridge between historical clinical descriptions of neural rigidity/overcontrol, in a subset of patients with JME and measurable fronto-temporal signatures across modalities.^13,40,41^

### Implications and limitations

Clinically, our findings underscore the importance of systematic assessment of psychiatric and personality pathology in JME. At the syndrome level, cognitive control and attentional vulnerabilities are common; within this landscape, psychiatric comorbidity appears to mark patients with more pronounced impairment, and OCPD identifies a subgroup with convergent medial prefrontal–cingulate and left fronto-temporal signatures that may be especially relevant under stress or arousal.

Several limitations merit emphasis. Subgroup sizes (particularly JME-OCPD in EEG and MRI) constrain power and may inflate effect-size estimates; replication in larger cohorts is essential. The cross-sectional design precludes causal inference regarding whether these markers precede epilepsy onset, reflect seizure/medication effects, or arise from shared developmental risk. Finally, our exploratory regression and correlation analyses were underpowered and relied on relatively coarse clinical predictors; more granular modelling of seizure dynamics, sleep, medication profiles, and stress reactivity, ideally within explicit active-inference formulations, will be important.

## Conclusion

Overall, the data support a neurodevelopmental account of JME characterized by executive–attentional dysfunction and altered fronto-temporal network organization. Within JME, comorbid OCPD is associated with a reproducible medial orbitofrontal–ACC thickening pattern and increased left fronto-temporal alpha coherence relative to psychiatrically unaffected JME patients, suggesting a distinct neurobiological phenotype plausibly linked to overcontrol and rigidity, particularly under challenge conditions.

## Disclosure of Conflict of Interests

Eugen Trinka reports personal fees from EVER Pharma, Marinus, Argenx, Arvelle/Angelini, Medtronic, Bial – Portela & Cª, S.A., NewBridge, GL Pharma, GlaxoSmithKline, Hikma, Boehringer Ingelheim, LivaNova, Eisai, UCB, Biogen, Genzyme Sanofi, GW Pharmaceuticals/Jazz, and Actavis outside the submitted work; his institution received grants from Biogen, UCB Pharma, Eisai, Red Bull, Merck, Bayer, the European Union, FWF Osterreichischer Fond zur Wissenschaftsforderung, Bundesministerium für Wissenschaft und Forschung, and Jubilaumsfond der Österreichischen Nationalbank outside the submitted work.

Georg Zimmermann gratefully acknowledges the support of the WISS 2025 project ‘IDA-Lab Salzburg’ (20204-WISS/225/197-2019 and 20102-F1901166-KZP)

The remaining authors have no conflicts of interest.

## Author contributions

LJR, BCP, JH, AT and ET contributed significantly to conception and design of the presented paper, acquisition, analysis and interpretation of the data as well as drafting of the paper.

ES, PL, KW, AT, LKr, SS-Y, AK, EL, MKr, GZ, HJ and ET contributed to acquisition and analysis of data and revising the paper for intellectual content.

GK, JH, and ET contributed significantly to conception of the study, interpretation of the results and gave final approval of the submitted version of the manuscript.

## Acknowledgements

This work was supported by the Austrian Science Fund (FWF), Project Number: KLI 543 B-27 and was generated within the European Reference Network for Rare and Complex Diseases EpiCARE.

## Data Availability

The data supporting the findings of this study are available from the corresponding author upon reasonable request. Study datasets are not made publicly available due to ethical and data protection restrictions.

## Ethical Publication Statement

We confirm that we have read the Journal’s position on issues involved in ethical publication and affirm that this report is consistent with those guidelines.

