## Supplement for "Neural correlates of Obsessive – Compulsive Personality Traits in Juvenile Myoclonic Epilepsy"

**Supplementary: Decoding the Janz Syndrome**

**Supplementary Methods**

S1. Neuropsychological and psychiatric assessment

S2. EEG modelling and connectivity measures

S3. MRI processing and surface metrics

**Supplementary Tables**

Table S1 (4): Pairwise neuropsychological group comparisons for JME with OCPD

Table S2: Multiple regression analyses of neuropsychological performance on OCPD status, seizure freedom, and ASM load

Table S3: Kendall’s tau correlations between alpha-band coherence and neuropsychological performance across all participants

Table S4: Attentional performance and executive functions in patients with JME and healthy controls

Table S5: Clusters that survived familywise-error (FWE) correction in group comparisons for cortical thickness

**References**

### Supplementary Methods

#### S1. Neuropsychological and psychiatric assessment

Test of Attentional Performance (TAP): Test of Attentional Performance (TAP; Version 2.2)[1] is a software package that can be used primarily to examine attentional and executive functioning. In our study, we used the subtests Alertness, Divided Attention and GoNogo in order to test for intrinsic and phasic alertness, shared auditory and visual attention, as well as inhibitory control, respectively.

Trail Making Test: The Trail Making Tests (TMT)[2,3] are neuropsychological instruments for detecting neurological disease and neuropsychological impairment in the domain of processing speed, sequencing, mental flexibility and visual-motor skills.

Wechsler Intelligence Scale: The Wechsler Intelligence Scale IV is an intelligence quotient test to measure various cognitive abilities in adults and adolescents.[4] We used the digit span (forward and backward) to measure the verbal attention span and working memory.

SCID I + II: The Structured Clinical Interview for Axis I disorders (SCID I) and for Axis II disorders (SCID II)[5], based on the Diagnostic and Statistical Manual of Mental Disorders-IV, is considered to be the gold standard for a semi-structured assessment of clinical disorders and personality disorders, respectively. German versions were used to assess DSM-IV Axis I and Axis II pathology.

#### S2. EEG modelling and connectivity measures

Based on findings of altered network connectivity in patients with JME[6] we analysed whether resting-state network connectivity in certain frequency bands differed between the groups and was furthermore correlated to certain differences in neuropsychological test scores. We focused on resting periods during hyperventilation, as these segments proved to be absent of artifact interference and epilepsy-related network disturbances are most likely to be recorded. Six patients had to be excluded due to poor signal quality or the lack of a hyperventilation period (PA11, PA14, PA27, PA31, PA47, & PA58). EEG analysis was performed using MATLAB (release R2023a, The Mathworks, Natick, MA, USA). EEG signals were re-referenced against an average reference channel based on the entire signal obtained from scalp electrodes excluding the recorded reference channel Cz as well as mastoid channels A1 and A2. Then we excluded all EEG channels, not relevant to our connectivity analysis. As we were interested in frontal and temporal channels only, the remaining channels were Fp1, F3,

F7, Fp2, F4, F8, T3, T5, T4, and T6. EEGs were then segmented into a period of 3 minutes starting with the hyperventilation initiation.

For each participant, autoregressive models were calculated using the mvfreqz.m and mvar.m function implemented within the BioSig toolbox[7] The multivariate autoregressive models were calculated for all channel x channel combinations and for the alpha frequency band (alpha: 8 – 13Hz). The model order was chosen individually for each subject in adherence with two boundary criterions: first, the maximum model order was set at 460, thus guaranteeing the adherence with a proposed ratio of 10:1 between given samples and the number of estimates.[8] Second, to estimate measures for 43 frequency components with a spatial sampling of 10 channels, the model order needed to be above 8.6 to ensure valid mapping of respective components.[9] Based on these restrictions, we performed a model optimization procedure, calculating for each subject and trial an autoregressive model with model orders 9 to 460 and deriving the Schwarz Bayesian criterion for each model using the artif2.m function of the tsa toolbox.[10] Using the so-defined optimal model order for each participant individually, we calculated multivariate autoregressive models.

From the results we derived real valued coherence, an undirected measure of connectivity, which is taking into account only the real part of the complex coherence[11] and which has been suggested to be a reliable (in terms of stability over time) marker in neurological conditions, such as epilepsy.[12] The measure was computed for 1 Hz frequency steps and then averaged according to the respective frequency band of interest. The resulting connectivity-matrices were then averaged according to the respective regions of interest: frontal left (Fp1, F3, F7), frontal right (Fp2, F4, F8), temporal left (T3, T5), and temporal right (T4, T6). Hence, for each subject we ended up with a 4 x 4 matrix of interactions for two derived connectivity measures in the alpha frequency band.

In the present study, to reduce the number of statistical tests and because our a priori hypotheses focused on alpha-band abnormalities in JME, only alpha-band coherence values were entered into group-level statistical analyses. For each subject we extracted six a priori defined connections between frontal and temporal ROIs (frontal left to frontal right, frontal right to frontal left, frontal left to temporal left, temporal left to frontal left, frontal right to temporal right, temporal right to temporal left). These alpha-band coherence values were then compared between groups defined by the OCD group variable (JME without psychiatric comorbidity, JME with any non-OCPD psychiatric diagnosis, JME with OCPD, and healthy controls). We performed Welch's t-tests (two-tailed, unequal variances assumed) for the following contrasts: (1) all JME patients vs. healthy controls, (2) JME with any psychiatric comorbidity vs. healthy controls, (3) JME with OCPD vs. healthy controls, and (4) JME with OCPD vs. JME without psychiatric comorbidity. Within each contrast, p-values were corrected for multiple comparisons across the six alpha-band connections using the Benjamini–Hochberg false discovery rate procedure; for each comparison we additionally quantified effect sizes using Hedges' *g*.

**S3. MRI processing and surface metrics**

Structural MRI data were acquired on a 3T Magnetom Prisma-Fit scanner (Siemens, Erlangen, Germany) using a 3D multiecho magnetization-prepared rapid gradient-echo (MPRAGE) sequence. Sequence parameters were: repetition time (TR) = 2.4 ms, echo time (TE) = 2.2 ms, inversion time (TI) = 1060 ms, flip angle = 8°, matrix size = 320 × 300, and isotropic voxel size = 0.8 × 0.8 × 0.8 $mm^3$. High-resolution T1-weighted images were visually inspected for artifacts and gross abnormalities; scans with severe motion or reconstruction artefacts were excluded prior to surface reconstruction.

Cortical surface reconstruction followed a standard automated pipeline using Freesurfer .[13] T1-weighted images were transformed into Talairach space, skull-stripped, and intensity-normalized. White and gray matter were segmented, and subject-specific white and pial surfaces were generated for each hemisphere, yielding surface meshes of approximately 160,000 vertices per hemisphere. All surfaces underwent visual quality control, and manual corrections of skull stripping and tissue boundaries were performed where necessary. Individual surfaces were then registered to a common template surface to allow vertexwise comparisons across participants.

To capture different aspects of cortical morphology, we focused on three surface-based markers: cortical thickness, white matter surface area, and local gyrification index (LGI). Cortical thickness was computed as the distance between corresponding vertices on the white matter and pial surfaces. Surface area at each vertex was defined as the average area of the six triangles adjoining that vertex on the white matter surface, indexing local cortical expansion or compression. LGI was calculated using an established surface-based implementation that quantifies, at each vertex, the amount of cortex buried within sulcal folds relative to the amount of visible cortex on the outer surface. Cortical thickness and surface area maps were smoothed with surface-based diffusion kernels of 20 mm full width at half maximum during preliminary analyses Because LGI computation already involves intrinsic smoothing, a smaller kernel of 5 mm was applied to LGI maps.

Vertexwise statistical analyses were conducted in BrainStat for MATLAB. In a first step, we modelled the effect of age on cortical thickness, surface area, and LGI in the healthy control group to identify the best-fitting age term for each metric (e.g., linear vs. non-linear effects). The resulting age model was then incorporated into mass-univariate general linear models for vertexwise group comparisons, with cortical thickness, surface area, or LGI as the dependent variable and group as the main factor of interest. All models were adjusted for age and sex; for surface area, total white matter volume was included as an additional covariate to account for global scaling effects. Statistical maps were thresholded using cluster-wise family-wise error correction ($p_{FWE} < 0.05$) with a cluster-defining threshold of $p < 0.025$, as implemented in BrainStat.[14] Cluster labels and peak coordinates were derived from the template surface for descriptive reporting.

### Supplementary Tables

Table S1. Pairwise neuropsychological group comparisons for JME with OCPD

| Measure | JME OCPD vs HC *t* | JME OCPD vs HC $p_{FDR}$ | JME OCPD vs HC Hedges *g* | JME OCPD vs JME non-OCPD *t* | JME OCPD vs JME non-OCPD $p_{FDR}$ | JME OCPD vs JME non-OCPD Hedges *g* |
|---|---|---|---|---|---|---|
| Digit span (back) | -1,16 | 0,355 | -0,41 | 0,18 | 0,862 | 0,07 |
| Digit span (fwd) | -1,37 | 0,25 | -0,38 | 0,43 | 0,672 | 0,12 |
| Inhibition (err) | 1 | 0,44 | 0,32 | 0,25 | 0,806 | 0,07 |
| Inhibition (rt) | 4,02 | 0,001 | 1,16 | 0,6 | 0,551 | 0,14 |
| Divided attention (vis) | 1,72 | 0,146 | 0,85 | 1,21 | 0,245 | 0,5 |
| Divided attention (aud) | 1,62 | 0,247 | 0,52 | 0,57 | 0,572 | 0,13 |
| Mental flexibility | 2,09 | 0,079 | 1,15 | 0,68 | 0,507 | 0,22 |
| Psychomotor speed | 3,13 | 0,011 | 1,44 | 1,02 | 0,323 | 0,34 |
| Tonic alertness (rt) | 3,43 | 0,006 | 1,64 | 1,21 | 0,235 | 0,31 |
| Phasic alertness (rt) | 2,25 | 0,056 | 1,05 | 0,75 | 0,463 | 0,21 |

*Note.* Welch's *t*-tests (two-tailed, unequal variances assumed) comparing patients with JME and OCPD to healthy controls (HC) and to JME patients without OCPD (JME non-OCPD). p(FDR) = p-values corrected for multiple comparisons across measures using the Benjamini–Hochberg false discovery rate procedure. Hedges' *g* indicates effect size (positive values reflect higher scores in the JME-OCPD group relative to the comparison group; negative values reflect lower scores). For reaction-time measures (inhibition RT, divided attention, psychomotor speed, tonic and phasic alertness), higher scores indicate slower performance.

Table S2. Multiple regression analyses of neuropsychological performance on OCPD status, seizure freedom, and ASM load

| Measure | n | *β* OCPD | $p_{FDR}$ | *β* seizure-free | $p_{FDR}$ | *β* ASM | $p_{FDR}$ | $R^2$ |
|---|---|---|---|---|---|---|---|---|
| TMT A | 41 | 6,24 | 0,578 | -2,44 | 0,887 | -7,85 | 0,329 | 0,155 |
| TMT B | 41 | 13,53 | 0,686 | -13,29 | 0,781 | 2,17 | 0,977 | 0,092 |
| Digit span (fwd) | 41 | -0,3 | 0,963 | 0,28 | 0,887 | 0,64 | 0,8 | 0,019 |
| Digit span (back) | 41 | -0,25 | 0,963 | 0,07 | 0,921 | 0,9 | 0,8 | 0,037 |
| Tonic alertness (rt) | 42 | 44,67 | 0,384 | -27,16 | 0,781 | -51,16 | 0,329 | 0,168 |
| Phasic alertness (rt) | 42 | 36,94 | 0,578 | -7,88 | 0,887 | -53,88 | 0,329 | 0,136 |
| Divided attention (aud) | 42 | 28,4 | 0,686 | -5,65 | 0,921 | -0,94 | 0,977 | 0,024 |
| Divided attention (vis) | 42 | 84,84 | 0,686 | -14,27 | 0,887 | 7,06 | 0,977 | 0,102 |
| Divided attention errors | 42 | 0,12 | 0,978 | -2,02 | 0,781 | 0,93 | 0,826 | 0,074 |
| Divided attention omissions | 42 | 1,23 | 0,578 | 0,44 | 0,887 | -0,76 | 0,8 | 0,056 |
| Inhibition (rt) | 42 | 16,13 | 0,716 | -8,8 | 0,887 | -32,45 | 0,502 | 0,048 |
| Inhibition (err) | 42 | -0,02 | 0,978 | -0,78 | 0,781 | 0,51 | 0,8 | 0,09 |

*Note.* Results of separate linear regression models for each neuropsychological outcome (one model per measure). Predictors were OCPD status (present vs. absent), seizure freedom (seizure-free vs. not seizure-free), and total antiseizure medication (ASM) load. *β* = unstandardized regression coefficient $p_{FDR}$ = *p*-value corrected for multiple comparisons across all 12 measures per predictor using the Benjamini–Hochberg false discovery rate procedure; $R^2$ = proportion of variance explained by the full model. No predictor remained statistically significant after FDR correction (all $p_{FDR} \geq .33$). Higher values in reaction-time measures indicate slower performance.

Table S3. Kendall's tau correlations between alpha-band coherence and neuropsychological performance across all participants

| Source | Sink | Cognitive measure | Kendall tau | $p$-value | $p_{FDR}$ |
| --- | --- | --- | --- | --- | --- |
| frontalleft | temporalright | GO_M | -0.183 | 0.012 | 1.000 |
| temporalright | temporalright | GO_M | -0.183 | 0.012 | 1.000 |
| frontalright | temporalright | TMT_B_RZ | -0.126 | 0.082 | 1.000 |
| frontalleft | frontalright | ZN_rw | -0.130 | 0.092 | 1.000 |
| frontalleft | frontalleft | VLMT_Lernleistung | -0.119 | 0.108 | 1.000 |
| frontalleft | frontalleft | Get_Fehler | -0.125 | 0.122 | 1.000 |
| frontalleft | temporalright | ZN_vw | -0.112 | 0.144 | 1.000 |
| frontalleft | temporalright | Get_Fehler | -0.113 | 0.161 | 1.000 |
| temporalright | temporalright | ZN_rw | 0.098 | 0.203 | 1.000 |
| temporalright | temporalright | ZN_vw | 0.097 | 0.205 | 1.000 |
| frontalright | temporalright | ZN_rw | 0.095 | 0.211 | 1.000 |
| temporalleft | temporalright | ZN_rw | 0.090 | 0.234 | 1.000 |
| temporalleft | temporalright | ZN_vw | 0.082 | 0.275 | 1.000 |
| frontalright | frontalright | ZN_vw | -0.081 | 0.279 | 1.000 |
| frontalright | frontalright | ZN_rw | -0.079 | 0.292 | 1.000 |
| temporalleft | temporalleft | ZN_rw | -0.074 | 0.322 | 1.000 |
| frontalright | frontalright | TMT_B_RZ | -0.067 | 0.363 | 1.000 |
| frontalleft | frontalright | ZN_vw | -0.063 | 0.393 | 1.000 |
| frontalright | frontalright | GO_M | -0.061 | 0.404 | 1.000 |
| temporalleft | temporalright | GO_M | -0.061 | 0.406 | 1.000 |
| temporalleft | temporalleft | GO_M | -0.060 | 0.414 | 1.000 |
| temporalright | temporalright | VLMT_Lernleistung | -0.056 | 0.440 | 1.000 |
| temporalright | temporalright | Get_Fehler | -0.055 | 0.448 | 1.000 |
| frontalright | temporalright | ZN_vw | -0.055 | 0.452 | 1.000 |
| frontalleft | frontalleft | ZN_vw | -0.052 | 0.468 | 1.000 |
| temporalleft | temporalleft | TMT_B_RZ | -0.051 | 0.478 | 1.000 |
| frontalleft | temporalright | TMT_B_RZ | -0.048 | 0.505 | 1.000 |
| frontalleft | frontalright | VLMT_Lernleistung | -0.046 | 0.515 | 1.000 |
| temporalleft | temporalleft | VLMT_Lernleistung | -0.044 | 0.529 | 1.000 |
| frontalright | temporalright | GO_M | -0.042 | 0.552 | 1.000 |
| temporalleft | temporalright | VLMT_Lernleistung | -0.038 | 0.580 | 1.000 |
| frontalright | frontalright | Get_Fehler | -0.036 | 0.607 | 1.000 |
| frontalleft | frontalright | Get_Fehler | -0.034 | 0.620 | 1.000 |
| frontalleft | frontalleft | TMT_B_RZ | -0.031 | 0.656 | 1.000 |
| frontalright | frontalright | ZN_vw | -0.030 | 0.664 | 1.000 |
| frontalright | frontalright | ZN_rw | -0.030 | 0.670 | 1.000 |
| frontalleft | temporalright | VLMT_Lernleistung | -0.028 | 0.684 | 1.000 |
| frontalleft | frontalright | TMT_B_RZ | -0.024 | 0.733 | 1.000 |
| temporalleft | temporalright | Get_Fehler | -0.023 | 0.744 | 1.000 |
| temporalleft | temporalright | TMT_B_RZ | -0.021 | 0.762 | 1.000 |
| frontalleft | temporalright | ZN_rw | -0.019 | 0.787 | 1.000 |
| temporalleft | temporalleft | ZN_vw | -0.018 | 0.795 | 1.000 |
| frontalleft | temporalright | GO_M | -0.014 | 0.834 | 1.000 |
| temporalleft | temporalleft | Get_Fehler | -0.013 | 0.848 | 1.000 |
| frontalright | temporalright | VLMT_Lernleistung | -0.012 | 0.854 | 1.000 |
| frontalright | temporalright | Get_Fehler | -0.011 | 0.867 | 1.000 |
| frontalright | frontalright | ZN_rw | -0.008 | 0.893 | 1.000 |
| frontalleft | frontalleft | GO_M | -0.007 | 0.902 | 1.000 |
| frontalright | frontalright | VLMT_Lernleistung | -0.006 | 0.906 | 1.000 |

| | | | | | |
|---|---|---|---|---|---|
| frontalright | temporalright | ZN_vw | -0.006 | 0.915 | 1.000 |
| frontalright | temporalright | Get_Fehler | -0.006 | 0.918 | 1.000 |
| temporalleft | temporalright | ZN_rw | -0.006 | 0.923 | 1.000 |
| frontalleft | frontalright | GO_M | -0.004 | 0.944 | 1.000 |
| temporalright | temporalright | TMT_B_RZ | -0.002 | 0.965 | 1.000 |
| temporalright | temporalright | GO_M | -0.002 | 0.972 | 1.000 |
| temporalright | temporalright | ZN_vw | 0.002 | 0.972 | 1.000 |
| frontalleft | frontalleft | ZN_rw | 0.005 | 0.946 | 1.000 |
| temporalleft | temporalleft | ZN_rw | 0.015 | 0.855 | 1.000 |
| temporalleft | temporalright | ZN_vw | 0.019 | 0.822 | 1.000 |
| frontalright | frontalright | TMT_B_RZ | 0.030 | 0.714 | 1.000 |
| frontalleft | frontalright | ZN_vw | 0.054 | 0.484 | 1.000 |
| frontalright | frontalright | ZN_vw | 0.084 | 0.292 | 1.000 |

*Note.* Kendall's tau rank correlations between alpha-band coherence for predefined EEG source–sink pairs and neuropsychological test scores (GO_M = Go/NoGo reaction time; TMT_B_RZ = Trail Making Test B; ZN_vw/ZN_rw = digit span forward/backward; VLMT_Lernleistung = verbal learning; Get_Fehler = divided-attention errors). Correlations were computed across all participants (JME patients and healthy controls combined). $p_{FDR}$ = *p*-values corrected for multiple comparisons across all 60 correlations using the Benjamini–Hochberg false discovery rate procedure. No correlation survived FDR correction (all $p_{FDR}$ = 1.00).

Table S4. Attentional performance and executive functions  in patients with JME and healthy controls

| | Control group | | JME patients | | Effect size | | Significance test | | | |
|---|---|---|---|---|---|---|---|---|---|---|
| | *n* | Median (IQR) | *n* | Median (IQR) | *RTE* | 95% CI | *t*-value | df | *p*-value | *p*-value adjusted |
| ***Attention*** | | | | | | | | | | |
| TMT-A (time in sec) | 67 | 22.00 (17.00 – 25.00) | 60 | 28.50 (22.00 – 34.25) | 0,725 | (0.635 - 0.815) | 4,968 | 107,697 | < 0.001 | **< 0.001** |
| TAP Tonic alertness (time in msec) | 66 | 229.50 (211.00 – 248.25) | 60 | 244.00 (221.75 – 275.25) | 0,652 | (0.554 - 0.749) | 3,071 | 106,858 | 0.003 | **0.036** |
| TAP Phasic alertness (time in msec) | 66 | 225.50 (215.25 – 241.50) | 60 | 235.00 (220.75 – 264.00) | 0,61 | (0.51 - 0.71) | 2,178 | 118,519 | 0.031 | 0.454 |
| TAP Divided - auditive (time in msec) | 66 | 619.50 (565.50 – 664.75) | 60 | 616.00 (555.25 – 691.00) | 0,537 | (0.433 - 0.641) | 0,707 | 114,395 | 0.481 | 1 |
| TAP Divided - visual (time in msec) | 66 | 783.50 (724.75 – 828.00) | 60 | 802.50 (746.25 – 859.50) | 0,577 | (0.475 - 0.678) | 1,495 | 121,779 | 0.137 | 1 |
| TAP Divided - Errors | 66 | 0.00 (0 – 1.00) | 60 | 1.50 (0 – 3.00) | 0,713 | (0.626 - 0.801) | 4,832 | 96,975 | < 0.001 | **< 0.001** |
| TAP Divided - Omissions | 66 | 1.00 (0 – 2.00) | 60 | 1.00 (0 – 3.00) | 0,559 | (0.459 - 0.66) | 1,171 | 105,729 | 0.244 | 1 |
| ***Executive Functions*** | | | | | | | | | | |
| TMT-B (time in sec) | 67 | 48.00 (41.00 – 59.50) | 60 | 60.00 (45.75 – 76.75) | 0,666 | (0.569 - 0.763) | 3,39 | 108,053 | < 0.001 | **0.016** |
| WIE Digit Span - forward | 67 | 11.00 (9.00 – 11.50) | 60 | 9.00 (7.75 – 11.00) | 0,374 | (0.276 - 0.472) | -2,552 | 117,833 | 0.012 | 0.231 |
| WIE Digit Span - backward | 67 | 7.00 (6.00 – 8.00) | 60 | 6.00 (5.00 – 7.00) | 0,367 | (0.27 - 0.463) | -2,732 | 120,764 | 0.007 | 0.185 |
| TAP GoNogo (time in msec) | 65 | 379.00 (346.00 – 418.00) | 60 | 413.00 (391.50 – 464.00) | 0,712 | (0.622 - 0.803) | 4,628 | 121,409 | < 0.001 | **0.001** |
| TAP GoNogo - Errors | 65 | 1.00 (0 – 2.00) | 60 | 1.00 (1.00 – 3.00) | 0,556 | (0.456 - 0.656) | 1,112 | 122,664 | 0.268 | 1 |

Abbreviations: TMT, Trail Making Test. TAP, testbattery for attentional performance. WIE, Wechsler Adult Intelligence Scale. NTrials, number of trials. TotCor, total number correct answers. TotErr, total number of errors. RTE, relative treatment effect. IQR, interquartile range.

Table S5: Clusters that survived familywise-error (FWE) correction in group comparisons for cortical thickness

JME with OCPD vs. healthy controls

| Cluster | Cohens *d* (95% CI) | $p_{FWE}$ | Regions |
|---|---|---|---|
| 1 | 0.83 (0.50, 1.4) | <.01 | Left medial orbitofrontal |
| 2 | 0.76 (0.51, 1.11) | <.05 | Left anterior cingulum |
| 3 | 0.73 (0.48, 0.98) | <.05 | Right lingual |
| 4 | 0.75 (0.49, 1.00) | <.05 | Right fusiform |
| 5 | 0.81 (0.50, 1.3) | <.01 | Right peri-calcarine |

JME without OCPD vs. heathy controls

| Cluster | Cohens *d* (95% CI) | $p_{FWE}$ | Regions |
|---|---|---|---|

No clusters survived multiple FWE error corrections.

JME with OCPD vs. JME without OCPD

| Cluster | Cohens *d* (95% CI) | $p_{FWE}$ | Regions |
|---|---|---|---|
| 1 | 0.98 (0.73, 1.23) | <.01 | Left medial orbitofrontal |
| 2 | 0.81 (0.51, 1.11) | <.05 | Left anterior cingulum |

**References Supplement**


1. Zimmermann P, Fimm B. *Testbatterie Zur Aufmerksamkeitsprüfung-Version 2.2:(TAP);[Handbuch]*. Psytest; 2009.

2. Bowie CR, Harvey PD. Administration and interpretation of the Trail Making Test. *Nature protocols*. 2006;1(5):2277-2281.

3. Reitan RM. Validity of the Trail Making Test as an indicator of organic brain damage. *Perceptual and motor skills*. 1958;8(3):271-276.

4. Drozdick LW, Raiford SE, Wahlstrom D, Weiss LG. The Wechsler Adult Intelligence Scale—Fourth Edition and the Wechsler Memory Scale—Fourth Edition. Published online 2018.

5. Wittchen HU, Zaudig M, Fydrich T. Skid. Strukturiertes klinisches Interview für DSM-IV. Achse I und II. Handanweisung. Published online 1997.

6. Caeyenberghs K, Powell HWR, Thomas RH, et al. Hyperconnectivity in juvenile myoclonic epilepsy: a network analysis. *NeuroImage: Clinical*. 2015;7:98-104.

7. Schlögl A, Brunner C. BioSig: a free and open source software library for BCI research. *Computer*. 2008;41(10):44-50.

8. Kus R, Kaminski M, Blinowska KJ. Determination of EEG activity propagation: pair-wise versus multichannel estimate. *IEEE transactions on Biomedical Engineering*. 2004;51(9):1501-1510.

9. Schlögl A, Supp G. Analyzing event-related EEG data with multivariate autoregressive parameters. *Progress in brain research*. 2006;159:135-147.

10. Schlögl A. Time series analysis toolbox. Published online 2010.

11. Nolte G, Bai O, Wheaton L, Mari Z, Vorbach S, Hallett M. Identifying true brain interaction from EEG data using the imaginary part of coherency. *Clinical neurophysiology*. 2004;115(10):2292-2307.

12. Höller Y, Butz K, Thomschewski A, et al. Reliability of EEG interactions differs between measures and is specific for neurological diseases. *Frontiers in human neuroscience*. 2017;11:350.

13. Fischl B, Dale AM. Measuring the thickness of the human cerebral cortex from magnetic resonance images. *Proc Natl Acad Sci USA*. 2000;97(20):11050-11055. doi:10.1073/pnas.200033797

14. Larivière S, Bayrak Ş, De Wael RV, et al. BrainStat: A toolbox for brain-wide statistics and multimodal feature associations. *NeuroImage*. 2023;266:119807.
